# Real-World Effectiveness and Safety of Psychedelic-Assisted Psychotherapy: Outcomes from a Large-Scale Compassionate Use Cohort in Switzerland

**DOI:** 10.1101/2025.12.01.25341335

**Authors:** T. Aboulafia-Brakha, A. Buchard, C. Mabilais, S. Alaux, C. Amberger, L. Furtado, F. Seragnoli, J-F Briefer, G. Thorens, M. Sabé, L. Szczesniak, R. Iuga, D. Zullino, L. Penzenstadler

## Abstract

**Background:** Classic serotonergic psychedelics such as LSD and psilocybin show promising antidepressant effects in controlled trials, but real-world data from routine clinical care remain limited.

**Methods:** This study retrospectively analysed routine data from adults with treatment-resistant depressive and/or anxiety disorders who received a first standardized Psychedelic-assisted Psychotherapy (PAP) cycle with 100 µg LSD or 25 mg psilocybin at a Swiss university hospital (May 2024–October 2025). Self-reported depression (BDI) and trait anxiety (STAI-T) were assessed at screening, one month before treatment, and 1–3 months post-treatment. In a subset of participants, cognitive emotion regulation (CERQ) was assessed pre- and post-treatment. Subjective drug effects and adverse events were recorded on the treatment day.

**Results:** The sample consisted of 115 patients (56.5% female; Mean age = 47.5 years). Depressive and anxiety symptoms significantly decreased over time (BDI: F(2,178) = 63.50, p < 0.001, partial η^2^ = 0.42; STAI-T: F(1.74,145.9) = 16.97, p < 0.001, partial η^2^ = 0.17), with no main effect of substance. CERQ analyses indicated reduced self-blame, rumination and catastrophizing, and increased positive refocusing and reappraisal. Perceived intensity followed distinct temporal profiles for LSD and psilocybin, but comparable subjective drug effects and clinical outcomes. Adverse events were mostly mild and transient, with no serious complications or treatment discontinuations.

**Conclusions:** In this compassionate-use real-world cohort, a first fully-active dose PAP session with LSD or psilocybin was well tolerated and associated with significant improvements in depressive and anxiety symptoms. These findings support the feasibility and effectiveness of PAP in specialised routine care.

## 1. Introduction

The therapeutic potential of Psychedelic-assisted Psychotherapy (PAP) for treating depressive anxiety disorders yielded a substantial increase in clinical research over the last decade [1-5]. To date, the pharmacokinetics and pharmacodynamics of various psychedelic substances are well-documented, and their safety and tolerability have been demonstrated in both healthy and clinical samples [3, 6-8]. Specifically, in patients with major depressive disorder (MDD), accumulating evidence indicates a significant and consistent reduction in depressive symptoms following one or two doses of classic serotonergic psychedelics (e.g., psilocybin, LSD-Lysergic acid diethylamide), often administered in combination with psychotherapy [9, 10]. Overall, the body of evidence is rapidly growing, with a focus on prospective controlled trials. While these prospective trials are undeniably essential in establishing safety and effectiveness, their highly constrained nature and selective patient populations can limit the generalizability of findings to real-world clinical practice.

Over the past decade, several phase 2 randomized controlled trials (RCTs) have shown that one or two sessions of psilocybin-assisted therapy can produce rapid, clinically meaningful reductions in depressive symptoms in patients with MDD [11, 12]. Recent systematic reviews and meta-analyses pooling these trials report at least moderate benefits of psilocybin-assisted therapy, while also highlighting small sample sizes, short follow-up and risks of unblinding and expectancy effects [13, 14]. Modern data on LSD are more limited but emerging. As such, a 2025 Swiss phase 2 trial found that two high-dose LSD sessions combined with supportive psychotherapy led to significantly greater and sustained improvements up to 3 months, in patients with moderate-to-severe MDD, with mostly transient adverse events [8]. While these emerging findings are promising, they also underline the need for further research, particularly studies examining how PAP can be implemented in routine clinical populations and real-world treatment settings. In Switzerland, the unique context of limited medical use of psychedelics (“compassionate use authorization”) provided by the Federal Office of Public Health (FOPH) under the Narcotics Law (LStup; RS 812.121) has created a significant opportunity for real-world clinical application. Over the last five years, the psychedelic-assisted psychotherapy (PAP) program at the Addiction Division, Department of Psychiatry of the Geneva University Hospital has been granted authorizations to treat with LSD or psilocybin more than 250 patients with treatment-resistant depressive or anxiety disorders. Routine clinical data, essential for symptom monitoring and assessment of treatment efficacy, have been systematically collected, with a structured data capture over the last 18 months. This systematic collection allows us to conduct the current retrospective study, aiming to share the procedures and clinical outcomes from this large patient cohort. By analysing this unique dataset, we expect to contribute to the understanding of real-world PAP applications, helping to bridge the gap between controlled research settings and the complexities of routine clinical care. We were particularly interested in and able to report on the following:

i) ***a***. The evolution of self-reported depressive and anxiety symptoms from initial screening to one to three months post-treatment with LSD or psilocybin, and ***b***. exploring associated changes in cognitive emotion regulation strategies from pre- to post-treatment.
ii) ***a***. Characterizing the kinetics of the intensity of perceived drug effects and, ***b***. the overall nature of the psychedelic experience.
iii) Assessing treatment safety and tolerability via reported adverse effects at the end of the treatment day.

## 2. Methods

This study retrospectively analyses patient data collected at our PAP program. The treatment procedures, as described in previous publications [15, 16], involve a series of clinical appointments and assessments before, during, and after the treatment day. In this retrospective analysis we included patients who had completed the first cycle of treatment and received 100 µg of LSD or 25 mg of psilocybin between May 2024 and October 2025 (since we had systematized and standardized the assessment procedure within this period^1^). Approval by the local ethics committee was obtained (ID: BASEC 2022-02015) and all patients had signed a general informed consent. The study was registered at clinicaltrial.gov (NCT: 07164287).

### 2.1 Patient’s inclusion and treatment procedures

#### 2.1.a Screening session and patient’s inclusion

Patients were referred to the program by their psychiatrists or psychotherapists. An initial screening evaluated each patient’s suitability for the program including a comprehensive review of their clinical and medical history, pre-existing psychiatric conditions, current and past medication use, and an examination for potential contraindications to PAP. Concurrently, the rationale, potential benefits, and risks of PAP were thoroughly discussed, and a complete treatment plan was developed. The choice of substance (either LSD or psilocybin) was based on both clinical symptomatology and patient preference. Patient preferences were often shaped by preconceived ideas about different psychedelics (e.g. information from peers, media, or previous experiences), as well as practical considerations such as cost (psilocybin being more expensive) and expected session duration. For example, patients with prominent anxiety about losing control sometimes opted for psilocybin because of its shorter-lasting psychoactive effects compared to LSD. Of note, the cost of the substance, CHF 128.40 per 100 μg of LSD and CHF 502.00 per 25 mg of psilocybin, is not currently reimbursed by health insurance. However, appointments and professional services were mostly covered.

The formal inclusion criteria for our program were: age 18 years or older; diagnosis of MDD or an anxiety disorder based on DSM-5 criteria [17]; and treatment resistance, defined as the absence of response to at least two different classes of antidepressants with adequate dose, duration, and adherence. Patients also had to be in ongoing psychotherapy and agree to temporarily discontinue specific psychotropic medications (e.g., mirtazapine, trazodone).

Exclusion criteria were diagnosis of psychotic or bipolar disorder, high suicidal risk, severe cardiovascular, hepatic, or neurological disorders affecting the central nervous system, and pregnancy or breastfeeding. For this retrospective study, our cohort consists of the patients who met these criteria during the study period. All treatment decisions were made under medical responsibility following a discussion in a multidisciplinary panel meeting composed by medical doctors, psychologists and nurses.

#### 2.1.b Regulatory Approval and Preparation

After initial assessment and signature of a clinical informed consent form, an application for the use of the psychedelic substance for the treatment of depression or anxiety was submitted to the federal public health authority (FOPH). Approval times varied between 3 to 6 months.

Once approval was granted, patients were scheduled for a preparation session, typically held 1 to 3 weeks before the planned treatment day. During this appointment, all aspects of the treatment day were reviewed, with a particular focus on optimising “set and setting,” including the autonomous choice of music and the option to bring personal objects as emotional anchors. Therapeutic goals and an explicit treatment “intention” were recalled from previous meetings; coping strategies for potential anxiety (e.g. breathing techniques, mindfulness exercises) were also introduced. Routine safety procedures included a blood test and electrocardiogram prior to the session to rule out unknown somatic conditions, and, for women of childbearing potential, a pregnancy test the day prior to treatment.

#### 2.1.c Treatment and Follow-up

The treatment day was scheduled to commence at 9:00 AM, with patients requested to arrive earlier for final preparations. Upon arrival, a brief medical check was performed, including an assessment of vital signs (heart rate, blood pressure, and body temperature) to ensure that the patient was in a suitable condition for drug administration. Any remaining questions or concerns from the patient were addressed at this time.

LSD was administered orally as a liquid solution (containing 0.1 mg/mL of LSD d-tartrate-certified solution for compassionate use from Biel, Switzerland), and psilocybin was given in capsule form (containing 5 mg of psilocybin dihydrate -certified for compassionate use from ReseaChem GmbH, Burgdorf, Switzerland), according to the individual treatment plan. The session took place in a quiet room at the outpatient clinic of the Division of Addictology, where patients were continuously accompanied and monitored by psychiatric nurses experienced in PAP; predefined safety procedures were available to manage any challenging reactions. After the acute drug effects had subsided, typically around 5:00 PM for psilocybin and 6:00 PM for LSD, a final medical assessment (including vital signs and mental status) was performed, and patients were discharged with instructions to avoid driving and accompanied by a friend or family member.

The day after the treatment, patients returned for an integration session to discuss their experience. The aim of this meeting was to embed insights from the psychedelic session into the ongoing psychotherapeutic work and daily life. In an open, non-directive conversation, patients described their subjective experiences (e.g. thoughts, emotions, imagery, bodily sensations), while the therapist helped link these to the previously defined treatment goals and to develop new perspectives. Together, concrete action plans were formulated, including strategies for dealing with anticipated challenges and ways to translate newly gained insights into behavioural change and longer-term improvement; patients then continued to be followed in their usual outpatient psychotherapy.

A follow-up session “amplification” was scheduled one to three months after the treatment. This session focused on evaluating the longer-term impact of the psychedelic experience on symptoms, functioning, and overall well-being, reflecting to which extent initial insights had been implemented in everyday life. Based on this joint review, the patient and treatment team decided whether an additional dosing session appeared clinically appropriate and, if so, refined or reformulated the therapeutic goals and subsequent dose for further treatment.

### 2.2 Outcome measures and assessments

The following questionnaires were systematically administered during the screening, preparation, and amplification sessions via Research Electronic Data Capture (REDCap):

- Beck Depression Inventory (BDI) [18]: This 21-item scale measures the severity of depressive symptoms, such as sadness, pessimism, and suicidal ideation. Scores on each item range from zero to three, with a higher total score indicating more severe depression.
- State-Trait Anxiety Inventory, Trait subscale (STAI-T) [19]: This self-report questionnaire assesses an individual’s stable anxiety traits, representing their habitual response to perceived stress. A higher total score on the STAI-T is indicative of a greater degree of anxiety.
- Cognitive Emotion Regulation Questionnaire (CERQ) [20]: This self-report questionnaire is used to identify specific cognitive strategies individuals employ to regulate their emotions. This questionnaire assesses various strategies, such as rumination, reappraisal, and perspective-taking.

Data related to acute treatment effects were collected during the treatment day. During psychedelic sessions, subjective effects were recorded at different time points. Patients were asked to self-rate the intensity of their experience at 30, 60, 90, 180, and 300 minutes after psychedelic intake on a scale ranging from 0 (none) to 10 (maximal intensity) [15]. Patients also completed the Mystical Experience Questionnaire (MEQ) [21, 22] to retrospectively rate their mystical experience at the end of the treatment day, which measures the intensity of the psychedelic experience through its global score and individual subscales. Adverse effects were also recorded using a standardized checklist.

### 2.3 Statistical Analysis

Beyond descriptive statistics, statistical analyses included repeated measures ANOVAs to examine changes in the BDI and STAI over time (screening, one month pre-treatment, and 1– 3 months post-treatment) and to compare outcomes between treatment groups (LSD, psilocybin). The analyses focused on the main effects of time and group as well as their interactions. To explore changes in strategies within the CERQ in a subset of participants (N= 67), we used a repeated-measures Multivariate Analysis of Variance (MANOVA) to test the pre-to-post intervention effect (Time) on the nine subscales, which were treated as dependent variables. A significant effect was followed up with univariate repeated-measures ANOVAs for each subscale to investigate specific changes. To analyse the perceived intensity of effects, a Generalized Estimating Equation (GEE) model was used to account for the repeated measures within each participant and to appropriately model the ordinal nature of the rating scale data. The perceived intensity ratings, measured on a 0–10 scale at five time points (30, 60, 90, 180, and 300 minutes post-drug intake), served as the dependent variable. The model included the main effects of molecule (LSD vs. psilocybin) and time, as well as their interaction (molecule x time). An independent working correlation matrix was specified to model the relationship between the repeated measures. To assess the significance of effects, Wald chi-square statistics were used. Adverse effects were treated as frequencies for each specific symptom, and we used chi-squared tests to compare adverse effects between groups. All statistical analyses were conducted using SPSS 28.0 software.

## 3. Results

### 3.1 Patient Demographics and Clinical Characteristics

The sample included 115 patients aged 20 to 77 years (M = 47.5, SD = 12.12), 56.5% of whom were female. Regarding educational attainment, 59.1% held an undergraduate degree or higher, 33% had completed secondary education or vocational training, and 6% had attained a lower secondary education degree.

Depressive disorder was identified as the primary diagnosis for 72% of patients and as a secondary diagnosis for 11.3%. Anxiety disorders were the primary diagnosis for 18.9% and a secondary diagnosis for 50%. Substance use disorder was the primary diagnosis for 8% of patients and a secondary diagnosis for 15.8%. Of note, personality disorder was recorded as a secondary diagnosis in 6% of the sample. However, as no standardized assessment for personality disorders was used, this figure is likely to underestimate their true prevalence.

Regarding antidepressant medication, patients were receiving treatment with Selective Serotonin Reuptake Inhibitors (SSRIs) (27.8%), including molecules such as Fluoxetine, Sertraline, Citalopram, Escitalopram, and Paroxetine. A further 20.9% were prescribed Serotonin-Norepinephrine Reuptake Inhibitors (SNRIs), including Venlafaxine and Duloxetine. Additionally, 11.3% were on antidepressants acting on other mechanisms (e.g., Bupropion, Mirtazapine, Trazodone, Vortioxetine); this latter group was required to taper off treatment prior to psychedelic administration. The remaining 40% were not currently on any antidepressant treatment, although they had been previously, as they met the criteria for treatment resistance. This subgroup was exclusively taking other medications, such as antipsychotics, anxiolytics, or hypnotics; antipsychotics were discontinued one week before treatment, anxiolytics and hypnotics were generally also discontinued., if necessary, this treatment could be used one day prior to treatment.

Seventy (70) patients received LSD and 45 psilocybin for whom data related to subjective effects and adverse effects were analyzed. For longitudinal analyses 57 patients receiving LSD and 34 receiving psilocybin completed self-reported assessment at the three time-points (screening, one month pre-treatment and one to three months post-treatment). Patients who had not completed assessment at these three time points were not included in the analysis (naturally excluded by repeated measures ANOVA’s as described below^2^). Indeed, six (5%) patients had not completed screening, 14 (12%) did not complete the one-month pre-treatment questionnaire, and six (12%) did not complete the post treatment assessment.

### 3.2 Self-reported depressive and anxiety symptoms

A repeated measures ANOVA on self-reported depressive symptoms **(Figure 1)**, as measured by the BDI, showed a significant main effect of time (F (2, 178) = 63.50, p < 0.001). The assumption of sphericity was met (Mauchly’s test p = 0.15). This effect was characterized by a large effect size (partial-eta = 0.42) and high achieved power (100%). We found no significant main effect of group (F (1, 89) = 0.012, p = 0.91) nor a significant interaction between group and time (F (2, 178) = 1.91, p = 0.15).

**Figure 1.**
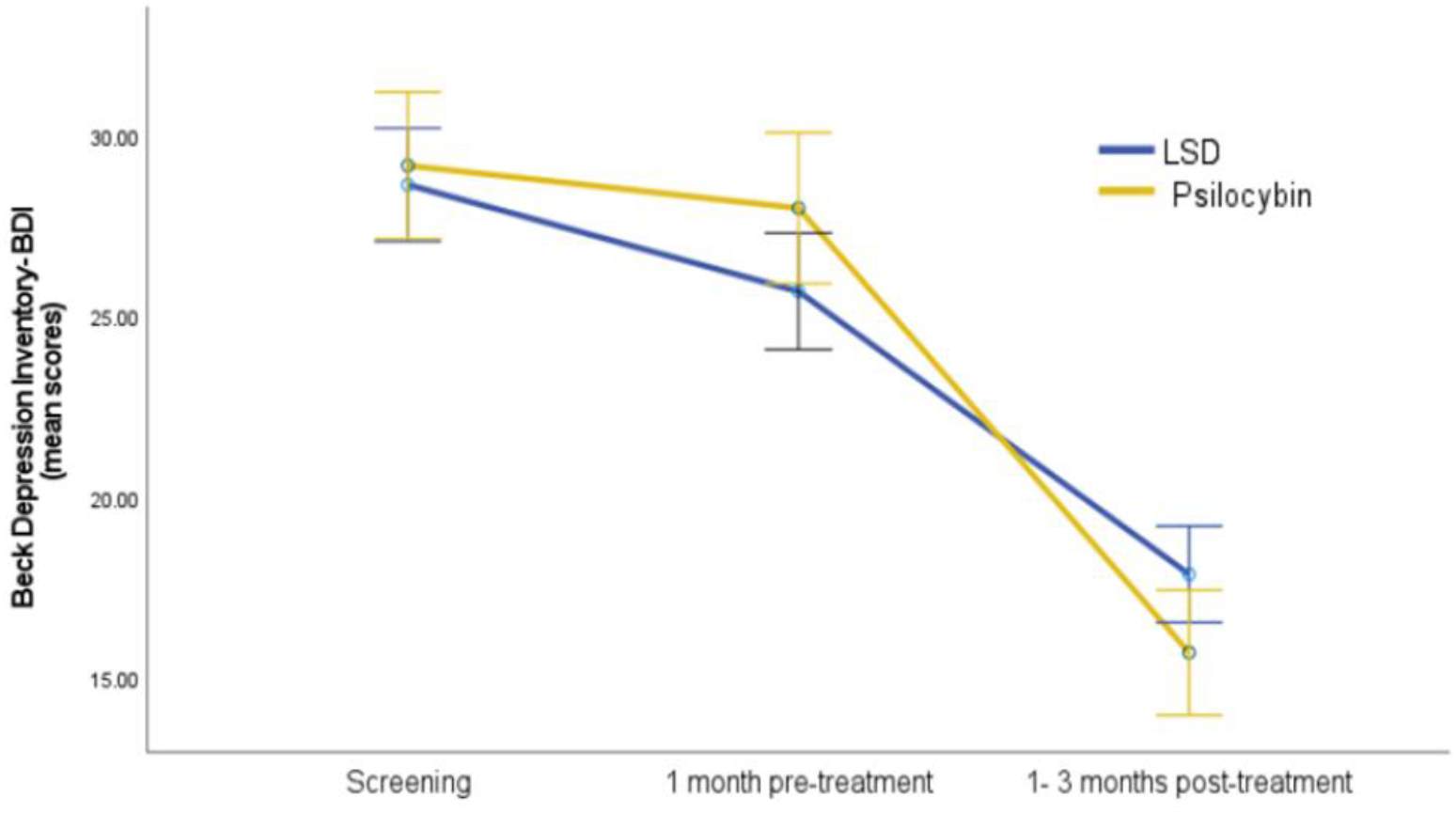
Self-reported depressive symptoms across time- Beck Depression Inventory. Error bars represent standard error from the mean.

The same pattern was observed for self-reported anxiety on STAI-trait scores **(Figure 2)**. As the assumption of sphericity was not met (Mauchly’s test p = 0.001), the Greenhouse-Geisser correction was applied. This analysis revealed a significant main effect of time (F (1.74, 145.9) = 16.97, p < 0.001), associated with a large effect size (partial-eta = 0.17) and high achieved power (99%). There was no significant main effect of group (F (1, 84) = 1.29, p = 0.26) or significant interaction (F (1.74, 145.9) = 0.14, p = 0.84).

**Figure 2.**
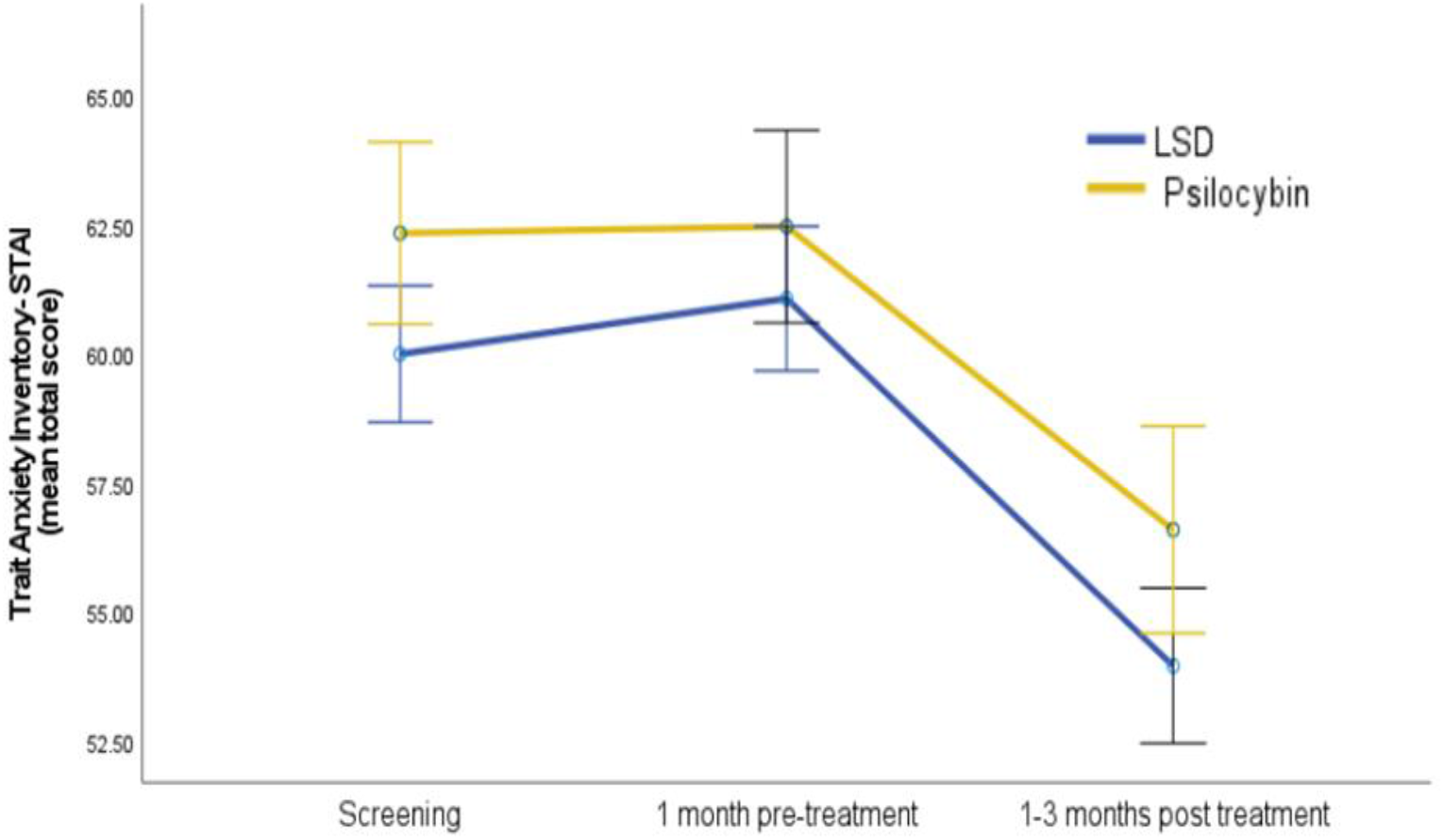
Self-reported anxiety symptoms across time- Trait anxiety inventory. Error bars represent standard error from the mean.

### 3.3 Cognitive emotion regulation strategies

The multivariate analysis on the combined set of CERQ subscores of a subset of the sample (receiving either LSD or psilocybin) revealed a statistically significant main effect of Time (from pre- to post-intervention) (F(9, 58) = 2.28, p = 0.028). Univariate follow-up analyses were then conducted for each subscale. Five subscales showed a significant change, with a decrease in Self-blame (p = 0.002), Rumination (p = 0.004) and Catastrophizing (p <0.001), associated with a significant increase in Positive Refocusing (p = 0.019) and Positive Reappraisal (p = 0.014). The subscale Putting into perspective showed only a trend to a significant increase (p= 0.063). The remaining subscales yielded no statistically significant changes: other-blame (p = 0.78), acceptance (p = 0.72) and Refocus on Planning (p = 0.24). Of note, following a Bonferroni correction given multiple comparisons, only a significant decrease in catastrophizing (p < 0.001), self-blame (p = 0.002), and rumination (p = 0.004) survived to strict significant threshold of p< 0.005.

### 3.4 Subjective effects

The GEE model revealed a significant main effect of time, confirming that perceived intensity changed over the course of the session (χ2=174.80, df = 4, p < 0.001) (Figure 3). However, there was no significant main effect of molecule on perceived intensity, suggesting that overall effects were similar for LSD and psilocybin (χ2=0.66, df = 1, p = 0.41). A significant interaction was observed between molecule and time (χ2=19.94, df = 4, p < 0.001). This shows that the two molecules had distinct temporal profiles, with their perceived intensity changing differently over the 5 time-points.

**Figure 3.**
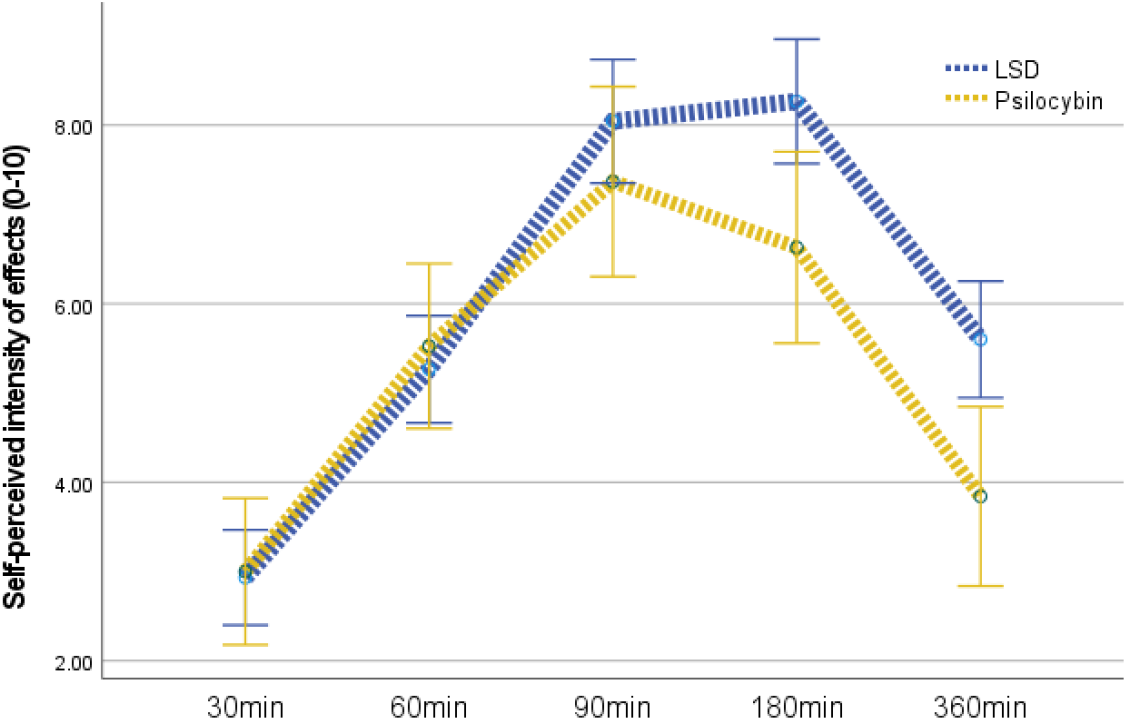
Self-perceived intensity of effects from 30 to 360 minutes after substance intake. Rating from 0 (no effects) to 10 (maximal effects). Bars represent standard error of the mean.

**Figure 4.**
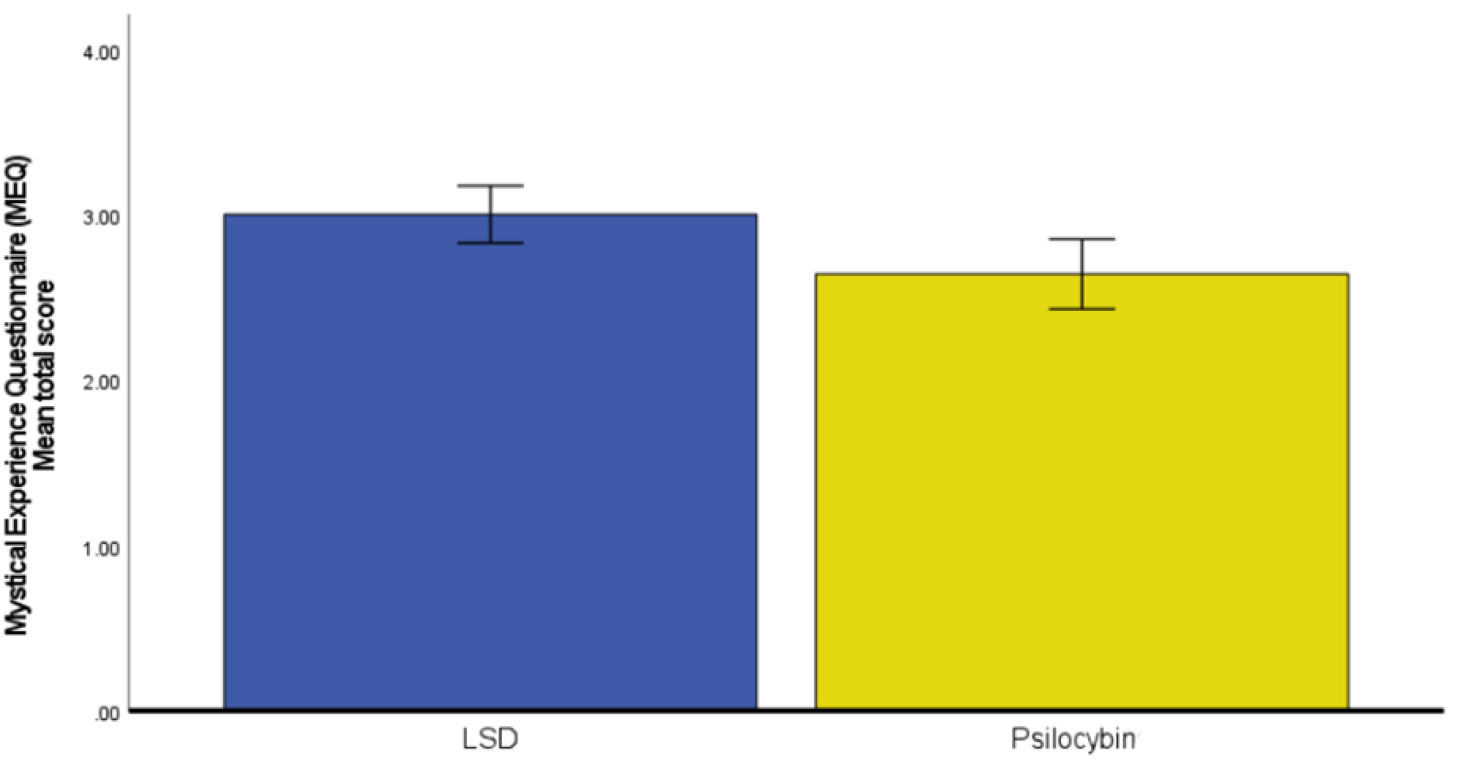
Total mean score at the mystical experience questionnaire (self-reported at the end of the psychedelic session on the same day). Error bars represent standard error of the mean.

There were no significant differences between the LSD and the psilocybin groups regarding self-reported intensity of the psychedelic experience as measured by global and subscale scores of the MEQ (F (1, 108) = 1.75, p= 0.19).

### 3.5 Adverse effects

Eighteen percent (18%) of patients who received LSD and 31.1% who received psilocybin reported no adverse effects. Table 1 shows the adverse effects reported by the two groups. There were no significant between-group differences regarding specific adverse effects. The most frequently reported events were blurred vision, dizziness, and nausea, with comparable frequencies observed for both the LSD and psilocybin groups.

**Table 1.** Adverse effects on the treatment day (percentage and number of patients reporting these effects)

| Adverse Effects | LSD<br>N= 70 | Psilocybin<br>N= 45 | Khi-squared, P<br>value |
| --- | --- | --- | --- |
| Blurred vision | 41.4% (29) | 35.6% (16) | 0.39, p= 0.53 |
| Dizziness | 31.4% (22) | 17.8% (8) | 2.65, p= 0.10 |
| Hot-cold<br>flashes/Chills | 28.6% (20) | 20% (9) | 1.07, p= 0.30 |
| Nausea | 24.3% (17) | 28.9% (13) | 0.30 p=0.58 |
| Tingling | 22.9% (16) | 22.2% (10) | 0.01, p= 0.93 |
| Drowsiness | 15.7% (11) | 24.4% (11) | 1.35, p= 0.24 |
| Headache | 12.9% (9) | 13.3% (6) | 0.005, p= 0.94 |
| Tremors | 11.4% (8) | 4.4% (2) | 1.68, p= 0.19 |
| Palpitations | 7.1% (5) | 4.4% (2) | 0.35, p=0.55 |
| Vomiting | 1.4% (1) | 0 | 0.65, p=0.42 |
| Tinnitus | 0 | 0 | - |
| Other | 14.3% (10) | 15.6% (7) | - |

## 4. Discussion

To the best of our knowledge, this is the first worldwide cohort study analysing real-world data from a compassionate use program using psychedelics for resistant depressive and anxiety disorders. The primary findings were a statistically significant and sustained reduction in self-reported depressive (BDI) and anxiety (STAI) symptoms over time. These results complement existing data from RCTs and suggest that psychedelic-assisted psychotherapy (PAP) may be effective in a population characterized by high treatment resistance, thereby strengthening the evidence for its external validity.

Of importance, symptom reduction was found to be independent of the specific psychedelic molecule administered. For instance, repeated measures ANOVAs revealed no significant difference between the LSD and psilocybin groups regarding clinical outcomes, nor a significant molecule-by-time interaction. This suggests that, for the primary therapeutic outcome in this heterogeneous population, the specific choice of the classic serotonergic psychedelic may be less critical than the overall therapeutic process. The present analysis focused exclusively on patients receiving a first standardized treatment cycle utilizing 100 µg LSD or 25 mg of psilocybin. The clinically meaningful changes observed with these doses align with the regimens used in current Phase 2 trials of psilocybin and LSD in depression, which generally involve one or two fully active-dose sessions [8, 13, 23]. It suggests therefore that a single, well-prepared high-dose session embedded in ongoing psychotherapy can produce substantial symptom improvements even in a broad, treatment-resistant population. Subsequent cycles were needed for some patients but were beyond the scope of this article.

The exploratory analysis of the (CERQ) provides an insight into specific changes in emotion related cognitive processing associated with the clinical improvement. The multivariate analysis of variance indicated a global shift in regulation strategies from pre- to post-treatment. The change primarily involved reduction in the three negatives strategies measured: catastrophizing, self-blame, and rumination and an improvement in positive refocusing and positive reappraisal. This pattern is consistent with process-based models that conceptualise depression as maintained by rigid, negative self-referential thinking and supports the notion that psychedelic-assisted treatment may act, in part, by enhancing psychological flexibility and loosening entrenched, self-critical cognitive styles. These findings converge with models positing that changes in psychological flexibility and emotion regulation may mediate reduction in depression and anxiety following psychedelic-assisted therapy [24-27]. Future prospective studies should explicitly test whether shifts in maladaptive cognitive-emotional strategies (e.g. reductions in rumination or self-blame) and changes in psychological flexibility mediate longer-term clinical outcomes after PAP, for example using pre–post assessments and longitudinal mediation models.

The overall intensity of the mystical experience was comparable between LSD and psilocybin. However, we observed a significant molecule by time interaction for perceived intensity, indicating distinct temporal profiles: LSD produced a more prolonged plateau, whereas psilocybin was characterised by a shorter overall duration with a similar peak intensity. This pattern is consistent with experimental pharmacology data from controlled cross-over studies in healthy participants [7, 28, 29]. Nonetheless, these kinetic differences do not seem to translate into detectable differences in short-term antidepressant or anxiolytic efficacy.

The adverse event profile observed in this cohort was in line with established safety data for classic serotonergic psychedelics, consisting mainly of mild, transient somatic and psychological reactions (e.g. blurred vision, dizziness, nausea), with no treatment discontinuations or molecule-specific differences. Rates and types of adverse events were comparable between LSD and psilocybin and closely mirror those reported in RCTs and experimental studies in both healthy volunteers and clinical populations [11, 23, 29, 30]. Taken together with the absence of serious or persisting complications in a sample characterised by high comorbidity and treatment resistance, these findings suggest that PAP can be delivered safely within a university hospital outpatient setting under a compassionate-use framework, provided that careful screening, close monitoring, and predefined crisis-management procedures are in place. More broadly, these real-world safety data from a relatively large cohort complement trial evidence and support the general tolerability of PAP outside strictly standardised research settings. Future studies should explore longer term effects.

### Limitations and future directions

The interpretation of these findings must consider the inherent limitations of a retrospective compassionate use study, including the lack of randomized control group, which prevents definitive causal attribution. Prospective designs with appropriate comparison groups (e.g. delayed-treatment or active-placebo conditions) are needed to better understand specific drug effects. In addition, patients in this program were referred, highly selected, and willing to wait several months for authorization and to undergo a demanding treatment day, which likely enriched the sample for motivation and psychological mindedness. Generalisability to less selected populations, other health systems, or settings with fewer psychotherapeutic resources therefore remains uncertain. Furthermore, the self-reported nature of clinical outcomes limits the robustness of our findings, as questionnaire data are vulnerable to recall bias, social desirability, and expectancy effects and were not complemented by standardized clinician-rated instruments. Future studies embedded in routine care should combine patient-reported outcomes with semi-structured clinical interviews and validated observer-rated scales to provide a more comprehensive and externally verifiable assessment of symptom change. In addition, the exploratory nature of the CERQ analyses necessitate cautious interpretation regarding changes in specific cognitive emotion regulation strategies. Future trials within clinical routine, should predefine mechanistic hypotheses and prioritise repeated assessments of cognitive–emotional processes, such as rumination, catastrophizing and other markers of psychological flexibility, as candidate mediators and clinical markers of change. This would allow formal mediation analyses to test whether improvements in maladaptive emotion regulation partly account for the antidepressant and anxiolytic effects of PAP.

In conclusion, this retrospective study, leveraging a large patient cohort treated under Switzerland’s compassionate-use framework, demonstrates an association between a first standardized PAP cycle (either 100 µg LSD or 25 mg psilocybin, embedded in ongoing psychotherapy) and significant reductions in depressive symptoms and trait anxiety within a treatment-resistant population. Importantly, no detectable differences were found between the two substances. Exploratory findings suggest concomitant shifts in cognitive emotion regulation, specifically a reduction in self-blame, rumination, and catastrophizing. Furthermore, the predominantly mild and transient adverse events support the feasibility of safely delivering compassionate use PAP in a university hospital outpatient setting. Controlled and mechanistic studies are still necessary to confirm objective effectiveness, clarify underlying causal pathways, inform the safe implementation of PAP in routine care, and assess longer term effects over many years.

*The authors declare no conflicts of interest (COI) relevant to the content of this article*.

## Data Availability

All data produced in the present study are available upon reasonable request to the authors

## Footnotes

1 Our program started progressively in 2020, while a standardized assessment from screening procedures to post-treatment was formally implemented in May 2024 with a computerized and anonymized database.

2 Of note, a sensitivity analysis using a Linear Mixed Model (accounting for missing data under the Missing at Random assumption) yielded comparable effects to the main analysis. We report the Repeated Measures ANOVA results here for parsimony and ease of interpretation.

## Notes

### Competing Interest Statement

The authors have declared no competing interest.

### Clinical Trial

NCT07164287

### Funding Statement

This study did not receive any funding

### Author Declarations

Approval by the Geneva local ethics committee Commission Cantonale d'Ethique de la Recherche sur l'être humain (CCER) was obtained (ID: BASEC 2022- 02015) and all patients had signed a general informed consent. The study was registered at clinical trial.gov (NCT:07164287).

